# Mechanically ventilated patients shed high titre live SARS-CoV2 for extended periods from both the upper and lower respiratory tract

**DOI:** 10.1101/2021.12.27.21268312

**Authors:** Zack Saud, Mark Ponsford, Kirsten Bentley, Jade M Cole, Manish Pandey, Stephen Jolles, Chris Fegan, Ian Humphreys, Matt P Wise, Richard Stanton

## Abstract

**Background:** SARS-CoV-2 infection can lead to severe acute respiratory distress syndrome needing intensive care admission and may lead to death. As a virus that transmits by respiratory droplets and aerosols, determining the duration of viable virus shedding from the respiratory tract is critical for patient prognosis, and informs infection control measures both within healthcare settings and the public domain.

**Methods:** We examined upper and lower airway respiratory secretions for both viral RNA and infectious virions in mechanically ventilated patients admitted to the intensive care unit of the University Hospital of Wales. Samples were taken from the oral cavity (saliva), oropharynx (sub-glottic aspirate), or lower respiratory tract (non-directed bronchoalveolar lavage (NBL) or bronchoalveolar lavage (BAL)) and analyzed by both qPCR and plaque assay.

**Results:** 117 samples were obtained from 25 patients. qPCR showed extremely high rates of positivity across all sample types, however live virus was far more common in saliva (68%) than in BAL/NBAL (32%). Average titres of live virus were higher in subglottic aspirates (4.5×10^7^) than in saliva (2.2×10^6^) or BAL/NBAL (8.5×10^6^), and reached >10^8^ PFU/ml in some samples. The longest duration of shedding was 98 days, while the majority of patients (14/25) shed live virus for 20 days or longer.

**Conclusions:** Intensive care unit patients infected with SARS-CoV-2 can shed high titres of virus both in the upper and lower respiratory tract, and tend to be prolonged shedders. This information is important for decision making around cohorting patients, de-escalation of PPE, and undertaking potential aerosol generating procedures.

**Summary:** Patients on intensive therapy infected with SARS-CoV-2 tend to be prolonged shedders, excreting virus for far beyond the time periods specified in current guidelines, and live virus titres can be extremely high in both the upper and lower respiratory tracts.

## INTRODUCTION

The Coronavirus disease-2019 (COVID-19) pandemic, caused by severe acute respiratory syndrome coronavirus 2 (SARS-CoV-2), has resulted in a global human death toll of 5.36 million (as of 21^st^ Dec, 2021) [1]. Common early disease symptoms include a dry cough, exertional shortness of breath, fatigue, lethargy, diarrhea and high-grade fever [2], and in 10-15% of cases, this can progress to severe pneumonia needing hospitalization. In 1-2% of cases the disease can lead to severe acute respiratory distress syndrome (ARDS) needing ICU admission and may lead to death [3]. As a virus that transmits by respiratory droplets and aerosols, determining the duration of viable virus shedding from the respiratory tract is critical for patient prognosis, and informs infection control measures both within healthcare settings and the public domain [4]. Symptoms may persist for weeks or even months post infection, however shedding of infectious viral particles almost never occurs beyond 10 days of symptom onset, even in hospitalized patients [5]. In a recent systematic review and meta-analysis including over 5000 individuals infected by SARS-COV-2 prior to June 2020, viral RNA was detectable up to 83 days in the upper respiratory tract; however, no study detected live virus beyond day 9 of illness [5]. Immune dysregulation is an important exception to this rule, with infectious virions recoverable from renal-transplant recipients and adults with humoral immunodeficiency months after symptom onset [6-9]. Individuals requiring admission to ICU are subject to both infection-mediated immune dysregulation [10] and iatrogenic immunosuppression [11]. We therefore hypothesized that adults with critically-ill COVID-19 may be susceptible to prolonged viral shedding and represent a nosocomial reservoir of infection. Furthermore, no study has investigated whether the 9 day ‘cutoff’ for live virus isolation applies to the lower respiratory tract or airways.

We therefore compared samples from critically-ill and recovering patients with COVID-19 for the presence of infectious SARS-CoV-2 virions. We examined a range of upper and lower airway respiratory secretions for both viral RNA and infectious virions in mechanically ventilated patients admitted to the ICU of the University Hospital of Wales, with samples from the oral cavity (saliva), oropharynx (sub-glottic aspirate), or lower respiratory tract (non-directed bronchoalveolar lavage (NBL) or bronchoalveolar lavage (BAL)) and compared this to qPCR for genomes. Together, we show infectious viral particles are readily recoverable from saliva and that critically ill mechanically ventilated patients can secrete extremely high levels of live SARS-CoV-2 from multiple sites in the respiratory tract well beyond the 20-day isolation period currently recommended by the CDC for patients with severe COVID-19 [12].

## METHODS

### Sample collection

All patients were consented prior to sampling. Saliva was collected using a cotton wool roll (Neutral Salivettes®, SARSTEDT, Numbrecht, Germany) which was left against the buccal mucosa for two minutes and then spun at 2000g to collect supernatant or washed through with Dulbecco’s Modified Eagle Medium if no supernatant was present after centrifugation. Subglottic endotracheal tubes are used in many ICUs as they reduce the incidence of ventilator associated pneumonia. Subglottic aspirates are removed every four hours in standard practice and discarded as waste. This fluid represents an accumulation of oropharyngeal secretions which accumulate under gravity above the endotracheal cuff. Broncholaveolar lavage was undertaken using a disposable Ambu® aScope™ 4 and Broncho Sampler Set (Ambu UK) with lavage of up to 80ml of sterile saline, alternatively a non-directed bronchoalveolar lavage was performed by inserting a suction catheter into the lung until resistance was met and 20ml of sterile saline inserted and slowly withdrawn. All the patients who were enrolled in this study received evidence-based treatment as per published health board or ICU directorate guidelines. Samples were secured in a biosecure box, and transferred to the BSL3 laboratory, then processed within 4 hours of collection. Patient baseline characteristics and administered treatments are listed in table 1 and supplementary data 1.

**Table 1.** Baseline Characteristics and Treatments of Patients

| Variable |  |
| --- | --- |
| Age in years, Median (IQR) | 59 (50 - 68) |
| Female sex, Number (%) | 9 (36%) |
| In-hospital mortality, Number (%) | 16 (64%) |
| Immunosuppression, Number (%) | 5 (20%) |
| Received Dexamethasone treatment, Number (%) | 23 (92%) |
| Received Remdesivir treatment, Number (%) | 15 (60%) |
| Received Tocilizumab treatment, Number (%) | 10 (40%) |

### Ethics statement

Sample collection (20th October to 8^Th^ December 2020) was undertaken as a service evaluation to see if samples could be processed from the respiratory tract and virus measured as an alternative to qRT-PCR. Subsequently, ethical approval was given via the COVID-19 ENLIST study (REC Reference 20/YH/0309; sponsor Cardiff and Vale University Health Board, REC board Yorkshire & The Humber - Bradford Leeds Research Ethics Committee).

### Plaque assays

All cells were grown in DMEM containing 10 % (v/v) FCS, and incubated at 37 °C in 5 % CO_2_. Plaque assays utilised Vero E6, a gift from the University of Glasgow/MRC Centre for Virology, UK. Virus was titrated onto Vero E6 cells transduced with Lentivirus vectors expressing ACE2 and TMPRSS2 and drug selected, to enhance virus entry [13]. 6-fold serial dilutions of each sample type were used to infect the Vero E6 ACE2/TMPRSS2 cells for 1 h at 37 □C with rocking. Following this, cells were overlaid with DMEM containing 2 % FCS, 1.2 % Avicel®, 50 ug/mL of Gentamycin (11491822, Fisher Scientific, UK) and 2.5 ug/mL of Amphotericin B (A2942, Sigma Aldrich, UK). After 72 h, the overlay was removed, and the monolayer washed and fixed with 100% methanol. Monolayers were stained with a solution of 25% (v/v) methanol and 0.5 % (w/v) Crystal Violet, then washed with water and plaques were enumerated.

### RNA extraction

100 uL of each sample was incubated with 10 uL of Proteinase K (19131, Qiagen UK) for 15 minutes at room temperature, after which the samples were placed in a water bath set to 70 □C and incubated for a further 15 minutes to inactivate the Proteinase K. 10 uL of RQ1 DNase buffer (M6101, Promega, UK) and 10 uL of RQ1 DNase (M6101, Promega, UK) was then added to the samples, which were subsequently incubated at 37 □C for 30 minutes. RNA was then extracted using the QIAmp Viral RNA Minikit (52904, Qiagen, UK), following the spin protocol, and eluting in 60 uL of Buffer AVE.

### qPCR assays

Quantitative RT-PCR testing for SARS-CoV-2 was carried out by amplification and detection of the E-gene using the following primers and probe; E_Sarbeco_F1 (ACAGGTACGTTAATAGTTAATAGCGT), E_Sarbeco_R2 (ATATTGCAGCAGTACGCACACA) and E_Sarbeco_P1 (FAM-ACACTAGCCATCCTTACTGCGCTTCG-BBQ). Gene copy load was quantified using an E-gene control plasmid (pEX-A128-nCoV_E_Sarbeco, Eurofins Genomics, Germany). Sample RNA extraction quality was assessed by detection of RNAse P transcripts [14] using the following primers and probe; RP-F (-AGATTTGGACCTGCGAGCG), RP-R (GAGCGGCTGTCTCCACAAGT) and RP-P (FAM-TTCTGACCTGAAGGCTCTGCGCG-BBQ). Reactions were carried out in 20 uL volumes containing; 4.4 uL QuantiTect Virus Mastermix (211013, Qiagen, UK), 0.2 uL QuantiTect Virus RT Mix (211013, Qiagen, UK), 0.4 uM forward primer, 0.4 uM reverse primer, 0.2 uM probe, 1 uL RNA extraction as template, 0.5 uL non-acetylated BSA (2 mg/mL, 6917, Sigma-Aldrich, UK), and the final volume was made up to 20 uL using nuclease-free water. RT-qPCR was conducted on a QuanStudio 3 instrument (ThermoFisher Scientific, UK) with the following cycle conditions; 50 □C for 20 minutes, 95 □C for 5 minutes, followed by 40 cycles of 95 □C for 15 seconds and 58 □C for 45 seconds (with recording).

### SARS-CoV2 variant identification

Variant analyses were carried out by sequencing a portion of the SARS-CoV-2 Spike protein gene using the following primers; S_Preamp_F (GTGTTAATCTTACAACCAGAACTCAATTAC) and S_Preamp_R (CACAGACTTTAATAACAACATTAGTAGCG). RT-PCR volumes and conditions were as stated above, with the exception of the annealing temperature being set to 55 □C as opposed to 58 □C. Sanger sequencing was outsourced to Eurofins Genomics, Germany, sequencing with the S_Preamp_F primer.

## RESULTS

A total of 117 samples (44 saliva, 32 subglottic, and 41 BAL) were obtained from 25 adults admitted to the ICU at the University Hospital for Wales, a tertiary referral center. All patients had a previously confirmed molecular diagnosis of SARS-CoV-2 based on nasopharyngeal swab, and none had received a SARS-CoV-2 vaccine at time of sampling. The median age of individuals included in the study was 59 years (range 37 to 76), with a male bias (16/25, 64%).

To determine whether levels of virus shedding differed at different sites within the respiratory tract, samples of NBAL/BAL, subglottic aspirate, and saliva, were taken from critically ill mechanically ventilated patients and assessed for both RNA genome levels, and the titre of live virus (figure 1). Consistent with a prior clinical and molecularly-confirmed diagnosis at admission, qPCR showed extremely high rates of positivity across all sample types (93 – 97%). In contrast, detection rates for live virus varied between sample types. The majority of saliva samples (30/44; 68%) contained live virus (figures 1 and 2), however this was not the case in subglottic aspirates and BAL/NBAL samples. Nevertheless, infectious virions were still detected in 14/32 (44%) subglottic aspirate samples and 13/41 (32%) BAL samples (figures 1 and 2).

**Figure 1:**
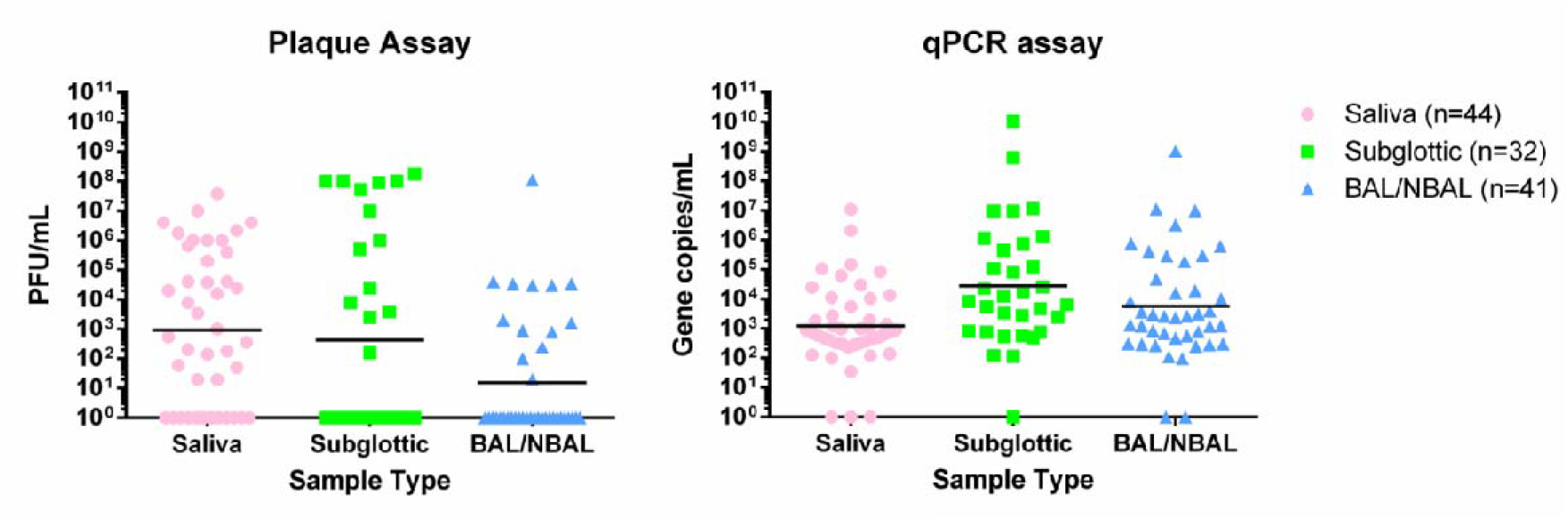
Titres of viable viral and gene copy load from saliva, subglottic aspirate, and bronchoalveolar lavage as determined by plaque and qPCR assays. Lines represent the geometric means.

**Figure 2:**
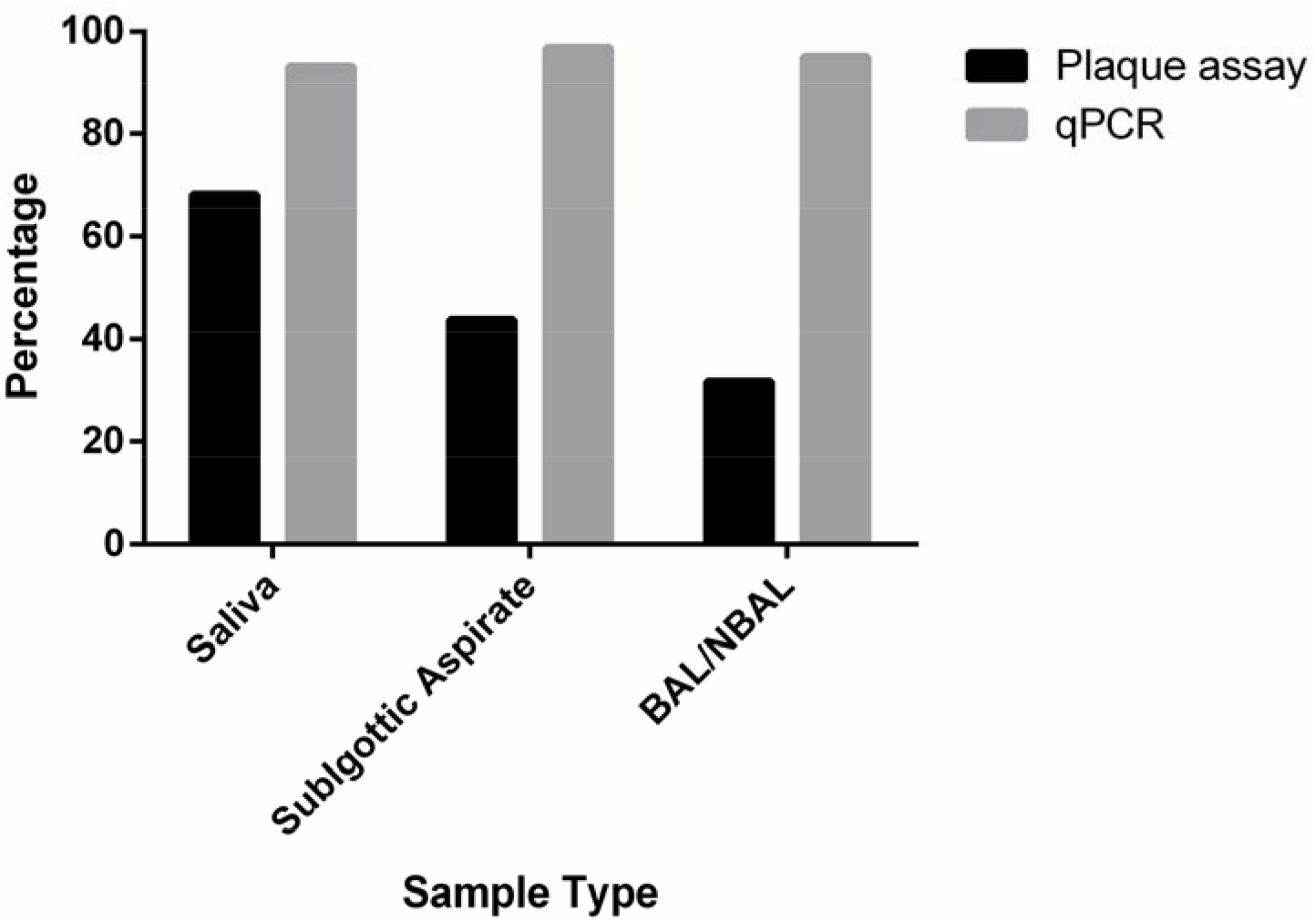
Percentage of all saliva, subglottic aspirate, and bronchoalveolar samples that were found to be positive by qPCR and plaque assays.

When titres of live virus were analysed, levels varied from the limit of detection (10 PFU/ml) to extremely high (>10^8^ PFU/ml). Across all samples, average titres largely reflected the chances of recovering live virus from any particular sample, with saliva containing the highest titre (1 × 10^3^ PFU/mL), while subglottic aspirates were slightly lower (2.5 × 10^2^ PFU/mL), and BAL lower still 1 × 10^1^ PFU/mL (Figure 1a). In contrast, when samples from which virus could not be isolated were excluded, subglottic aspirates contained significantly higher titres of live virus (4.5 × 10^7^ PFU/mL) than either saliva (2.2 × 10^6^ PFU/mL) or BAL/NBAL (8.5 × 10^6^ PFU/mL). This latter result was also reflected in the analysis of genome copy number, which was notably higher in subglottic aspirates than in saliva or BAL.

Previous studies have demonstrated a strong correlation between Ct value by qPCR and the chances of recovering live virus from oral swabs, with isolation of live virus becoming much more infrequent as Ct values increase. In accordance with this, qPCR was clearly more sensitive than virus isolation in saliva, BAL/NBAL, and subglottic aspirates. However, when virus titres were compared with genome titres, we did not observe strong correlations (figure 3). In saliva, samples lacking live virus all had genome titres below 10^4^ copies/ml, suggesting a ‘cut-off’ for detection of infectious virus. However, amongst samples containing live virus, genome titres were as low as 10^2^ PFU/ml. The correlation was even weaker in BAL and subglottic aspirates, where we failed to isolate live virus from samples containing RNA levels as high as 10^9^ genomes/ml, but successfully isolated virus from samples with genome titres of 10^3^ genomes/ml.

**Figure 3:**
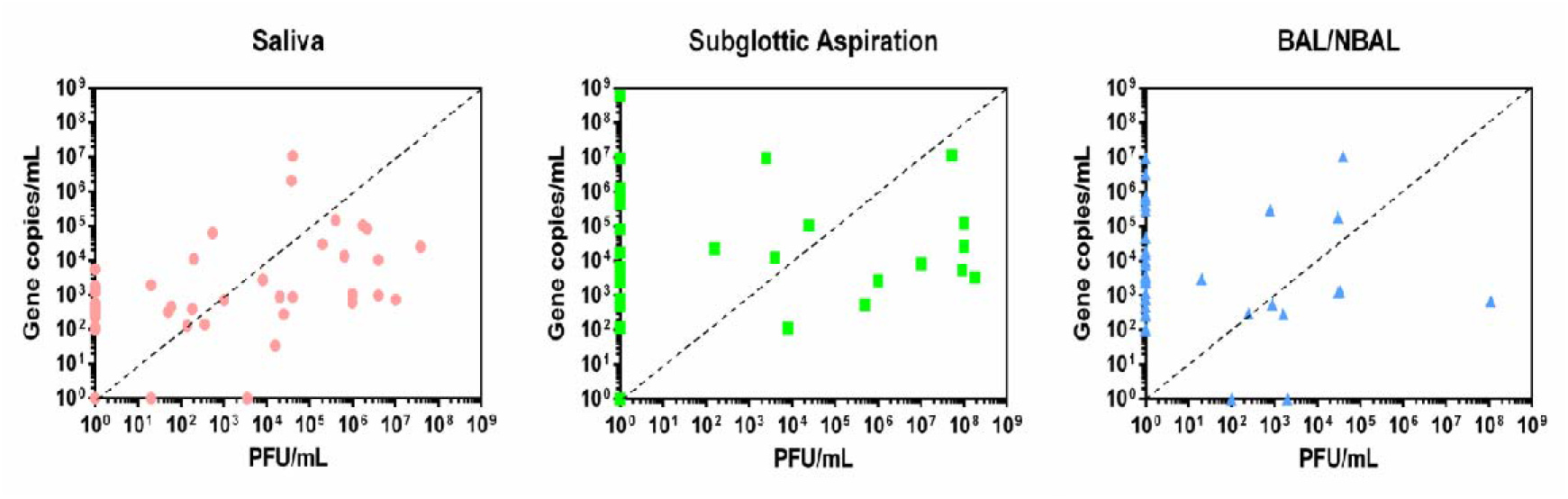
Correlation between viral load as determined by plaque assay (PFU/mL) and gene copies / mL as determined by qPCR assay. Comparisons were made between the saliva, subglottic aspirations, and BAL/NBAL sample types. The dashed line represents equal titres of the gene copy and viable viral loads.

Previous data suggests that, even amongst hospitalized patients, live virus is rarely detected beyond 10 days after symptom onset from oro-or nasopharyngeal swab samples. The situation was markedly different in our cohort, where 16 of the 25 patients shed viable virus for longer than 10 days (figure 4, supplementary data 1). The longest duration of shedding was 98 days, while the majority of patients (14/25) shed virus for 20 days or longer. When grouped by patient, saliva and subglottic aspirate tended to remain positive for longer than BAL, in accordance with our previous observation that BAL was the sample least likely to contain viable virus.

**Figure 4:**
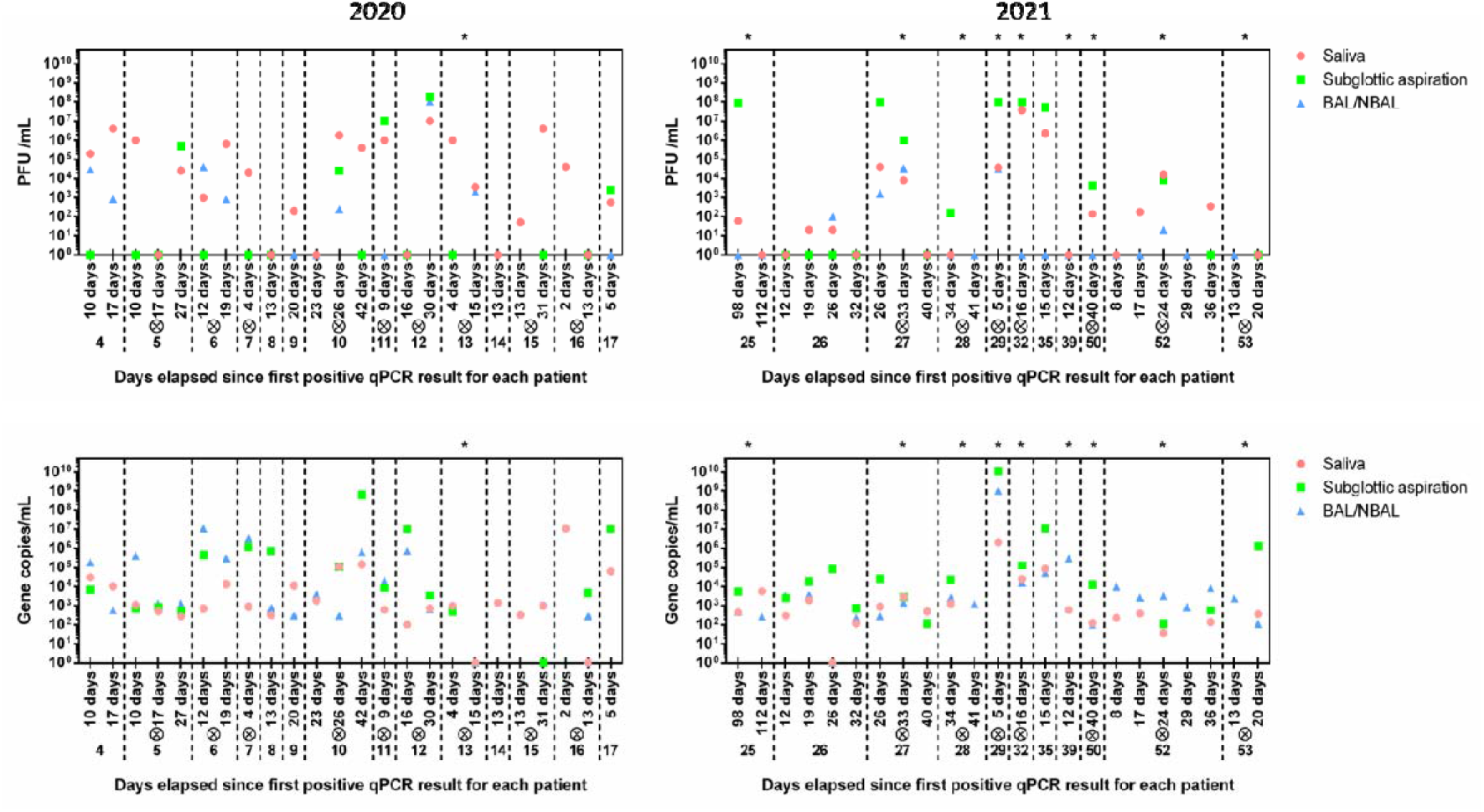
Longitudinal analyses of patient viral and gene copy loads as determined by plaque and qPCR assays, respectively. A cross inside a circle above the patient number indicates a fatal outcome for the patient. Asterisks above a patient block indicate the patient was infected with the Alpha variant (B.1.1.7).

While this study was underway, the Alpha VOC began to spread. We therefore sequenced the Spike gene to determine which variant each patient was infected with, to determine whether the isolation of viable virus differed based on strain. No clear differences were seen in the longevity of virus isolation. Furthermore, no correlation was observed between viral load and patient outcome.

When viable viral and gene copy loads from each patient were compared longitudinally, the highest viral titres across all patients were recorded from subglottic aspiration samples. However overall, saliva provided a better indication of infection; whenever live virus was isolated from any sample at any timepoint, saliva at that timepoint always contained live virus. In contrast, by qPCR, sample type was largely irrelevant for determining positivity. There were however differences in viral load by qPCR, with subglottic aspirates often containing higher titres than saliva.

## DISCUSSION

Current NHS guidance state isolation precautions can be discontinued in most individuals with SARS-CoV-2 infection 10 days after symptom onset [15], whilst the Centers for Disease Control and Prevention (CDC) recommends extending this for up to 20 days after symptom onset in those with severe illness [12]. Our study clearly demonstrates that ICU patients can frequently excrete high titres of infectious SARS-CoV2 for periods far exceeding these recommendations. Viral titres from saliva, subglottic aspirate, and BAL/NBAL can reach titres of >10^7^ PFU/ml in some patients. Furthermore, these levels of virus did not appear to be variant specific as individual patients infected by either Alpha, or earlier variants, shed these high titers of infective virus. Our study also highlights the inadequacy of qPCR in determining the point during the infection course when an intubated patient ceases to present an infection risk to hospital staff; this was particularly true for samples from the airway and lower respiratory tract, where PCR positivity was poor at predicting the presence of live virus. Furthermore, in contrast to studies using oral swabs in hospitalized patients [25], Ct value from qPCR of ICU patients was not a good predictor for the presence of live virus, a problem that has been highlighted in previous studies investigating discrepancies between RT-PCR results and symptomatic infection [26-28].

Only one other study has titrated live virus from clinical samples, demonstrating titres of 5×10^6^ and 4×10^6^ PFU/ml in nasopharyngeal swabs from two patients [18]. Thus, despite their prolonged shedding, titres in ICU patients are not dramatically higher than those in people with milder disease. Mouth swabs or saliva are commonly used to diagnose SARS-CoV-2 infection, and may be interpreted as a surrogate for shedding of live virus. However, the respiratory droplets that transmit virus have been assumed to arise from both the upper and lower respiratory tract. Previous studies have used molecular methods to compare viral genome loads in BAL compared to mouth swabs [16, 17]. In agreement with these studies, we find that PCR results are largely concordant between upper and lower respiratory tract samples, thus BAL/NBAL samples do not offer an advantage over the more practical saliva or nasopharyngeal samples for diagnosis. However, in contrast to viral genome, live virus was much more common in saliva than BAL or NBAL, suggesting that the upper respiratory tract is more likely to be a source of infectious virus than the lower. This is consistent with previous reports that there is independent replication of virus in the upper and lower airways [29]. However, it may also reflect the chances of virus being inactivated in a sample containing high levels of mucus and other proteolytic enzymes, and the volume of fluid used to lavage the lungs parenchymas. Titres in BAL/NBAL may therefore be an under-estimate of the true situation. Nevertheless, when virus was present, titres were similarly high to other sites, frequently reaching >10^5^ PFU/ml. Thus, it is clear that cell free live virus is capable of reaching extremely high titres in the lungs, and is a potential source of transmissible virus in a proportion of patients. This discordance between titres of infectious virus and viral genomes in the BAL/NBAL sample highlights the advantage of measuring infectious viral load directly by plaque assay. Utilizing more commonly implemented indirect viral load measurement methods, such as inferring viral viral load by measuring gene copies and subsequently confirming the sample to contain infectious virions by observing CPE on cultured cells [30-32], would have resulted in drastic over and under estimation of infectious viral load in numerous tested samples.

In a proportion of samples, the genome titres were lower than the titres for live virus. This likely reflects the difficulty of extracting RNA from a highly proteolytic sample, and the need to process the sample to extract RNA in the absence of carry-through inhibitors – problems which are reduced in nasopharyngeal swabs that most studies use. All qPCR reactions were controlled by amplifying RNaseP, to ensure that PCR inhibitors did not affect results, and this is reflected in the fact that nearly all samples were positive for viral RNA. Nevertheless, the higher processing requirements, and the fact that RNA is highly labile, may result in the genome copy number being an under-representation of the ***in vivo*** situation. Despite this, the gene copy load in our cohort was similar to those previously reported from oropharyngeal swabs [25] and saliva [33] samples in hospitalized patients.

Our study demonstrates that qPCR is not a robust indicator of viable viral shedding in critically ill patients, irrespective of the sample type. Patients on intensive therapy infected with SARS-CoV-2 tend to be prolonged shedders, excreting virus for far beyond the time periods specified in current guidelines, and live virus titres can be extremely high in both the upper and lower respiratory tracts. This information is important for decision making around cohorting patients, de-escalation of PPE, and undertaking potential aerosol generating procedures, more so given the threat of new variants, such as Omicron, that have higher transmission rates and greater vaccine escape potential. It also supports the continued use of oral antiseptics in these patients; products such as chlorhexidine are used routinely to reduce the incidence of ventilator-associated pneumonia [34]. Our data suggests that they may also have a role to play in minimising nosocomial transmission, although formulations containing surfactants are likely to be more effective in this role than chlorhexidine [35]. Future studies will be needed to assess whether the use of monoclonal antibody therapies, vaccination, and antivirals can reduce persistent shedding. Our study also highlights the need for more robust, practical assays for the determination of viable viral shed in healthcare settings.

## Data Availability

All data produced in the present work are contained in the manuscript

## ACKNOWLEDGEMENT

This work was supported by funding from the MRC (MR/V028448/1, MR/S00971X/1), Wellcome Trust (204870/Z/16/Z), Welsh Government Ser Cymru Scheme, and Accelerate Wales.

## Supplementary data

**1a**. Individual patient baseline characteristics and treatment regimen.

**1b**. Individual sample viral load, gene copy load, variant sequence, RNAseP Ct, and days exhibiting positive assay result.

**1c**. Individual sample qPCR Ct values and gene copy load calculations.

